# JADE: Jawbone Lesion Diagnosis and Decision Supporting System

**DOI:** 10.64898/2026.01.26.26344704

**Authors:** Soroush Baseri Saadi, Jonas Ver Berne, Rocharles Cavalcante Fontenele, Peter Claes, Reinhilde Jacobs

## Abstract

**Objectives:** To develop and evaluate JADE, a proof-of-concept retrieval-augmented generation (RAG) diagnostic assistive system was designed to enhance large language model (LLM) reasoning for the assessment of jawbone lesions. This study examined whether integrating structured retrieval with GPT-5 improves diagnostic accuracy and stability compared with standalone LLMs.

**Methods:** JADE was developed as a cloud-based application integrating GPT-5 with a curated oral radiology and pathology database using a hybrid semantic-keyword retrieval strategy. Clinical and radiographic characteristics were imported as a structured query to guide retrieval and support diagnostic reasoning. Performance was compared with standalone GPT-5, Claude Sonnet 4.5, DeepSeek-R1, and Gemini 2.5 Flash across 25 cases. Accuracy was analysed using Cochran’s Q test with post-hoc McNemar’s tests and Bonferroni correction. Intra-model stability was measured using the majority agreement ratio, and response time was recorded to assess real-time usability.

**Results:** JADE showed the highest diagnostic performance, correctly identifying 20 out of 25 cases and outperforming all standalone LLMs. Significant differences were observed across models (Cochran’s Q = 33.2, df = 4, *p* < 0.001), with post-hoc analyses confirming that JADE significantly outperformed GPT-5, Gemini 2.5 Flash, and Claude Sonnet 4.5 (*p* < 0.01). JADE also exhibited the greatest run-to-run stability (mean MAR = 0.90 ± 0.18). The average prediction time of 6 ± 0.5 seconds supported its feasibility for real-time clinical use.

**Conclusions:** JADE improved diagnostic accuracy and stability over standalone LLMs, underscoring the value of RAG reasoning in jawbone lesion assessment and its potential for real-time clinical use.

## Introduction

Diagnosis of jawbone lesions is a highly challenging task in clinical practice, as these lesions have diverse origins, unique anatomical structures, and often overlapping radiographic characteristics ^1^. Even though some lesions may be treated similarly, ranging from medication to surgical intervention, accurate differentiation and diagnosis are necessary to avoid unnecessary invasive procedures or inappropriate medical treatments ^2^.

Panoramic radiography is commonly used in daily dental practice as the main imaging modality for diagnosis, as it provides a broad two-dimensional view of the dentomaxillofacial structures ^3^. However, interpreting panoramic radiographs is quite a complex task, especially for a general dentist who often lacks the knowledge of an oral radiologist. Besides, considering the global shortage of oral radiologists, the diagnostic process can be time-consuming and subject to error.

Artificial intelligence (AI) has demonstrated great potential in medical diagnosis in oral healthcare. Deep learning (DL), a subfield of AI, in combination with image data, has shown promising results in the detection and classification of jawbone lesions on panoramic radiographs ^4,5^. However, precise diagnosis of jawbone lesions is beyond image analysis and requires the incorporation of multiple clinical and radiographic parameters.

Large language models (LLMs), as text-based generative AI systems, have strong contextual abilities, allowing them to interpret various clinical parameters and generate clinically relevant diagnostic reasoning. Advanced LLMs, such as OpenAI’s GPT series^6–8^, DeepSeek ^8,9^, and Gemini ^10,11^, have shown their abilities in providing diagnostic support and improving accuracy in oral pathology. Despite these advancements, the responses of LLMs are limited by their static training knowledge and the lack of deep domain-specific expertise. As a result, they may generate fluent but inaccurate or fabricated statements, known as hallucinations ^12–15^. Such outdated or generalised knowledge restricts their reliability as a clinical decision-support tool and highlights a critical gap that limits their effective use in oral pathology diagnosis.

To date, no study has investigated whether the diagnostic reasoning power of LLMs in oral pathology can be improved by extending their knowledge with domain-specific information beyond their general training. In particular, the potential of the retrieval-augmented generation (RAG) framework ^16^, which can enhance LLMs’ reasoning through external and up-to-date information, has not been explored for the diagnostic interpretation of jawbone lesions.

Therefore, this proof-of-concept study aimed to develop and evaluate JADE, a novel RAG-based system designed as a cloud-based, mobile-adapted application to support the differential diagnosis of jawbone lesions. This study examined whether this RAG-based system improves diagnostic accuracy over its underlying standalone LLM (GPT-5) and further compared its diagnostic performance and run-to-run stability with other standalone LLMs.

## Methods

### Study design

This study was divided into two main parts. The first part describes the architecture of the proposed application, explaining the components and workflow of the system. The second part focuses on the validation procedure, where the performance of the system was evaluated using a retrospective validation dataset to assess its diagnostic accuracy and clinical applicability in identifying jawbone lesions.

### Ethical considerations

This study, with a retrospective validation dataset, was carried out at the Centre of Dentomaxillofacial Radiology and the Department of Oral and Maxillofacial Surgery, University Hospitals Leuven, Leuven, Belgium. The Institutional Ethics Committee approved the study protocol (approval number: S65708), and all procedures adhered to the World Medical Association Declaration of Helsinki and the Institutional Review Board. Patient data were anonymised before analysis to ensure data protection and confidentiality. Moreover, no personal identifiers or metadata were used in this study beyond the radiographic and diagnostic information.

## Application architecture

JADE, both with and without its RAG component, is a cloud-based diagnostic support application designed for mobile and tablet platforms, providing a user-friendly interface for general dentists, oral radiologists, and dental students. It functions as an intelligent system for the diagnosis of jawbone lesions by integrating clinical data and radiographic characteristics of lesions into a structured diagnostic workflow. Based on this input, the system automatically generates the most likely differential diagnosis for the described jawbone lesion (Figure 1).

**Figure 1.**
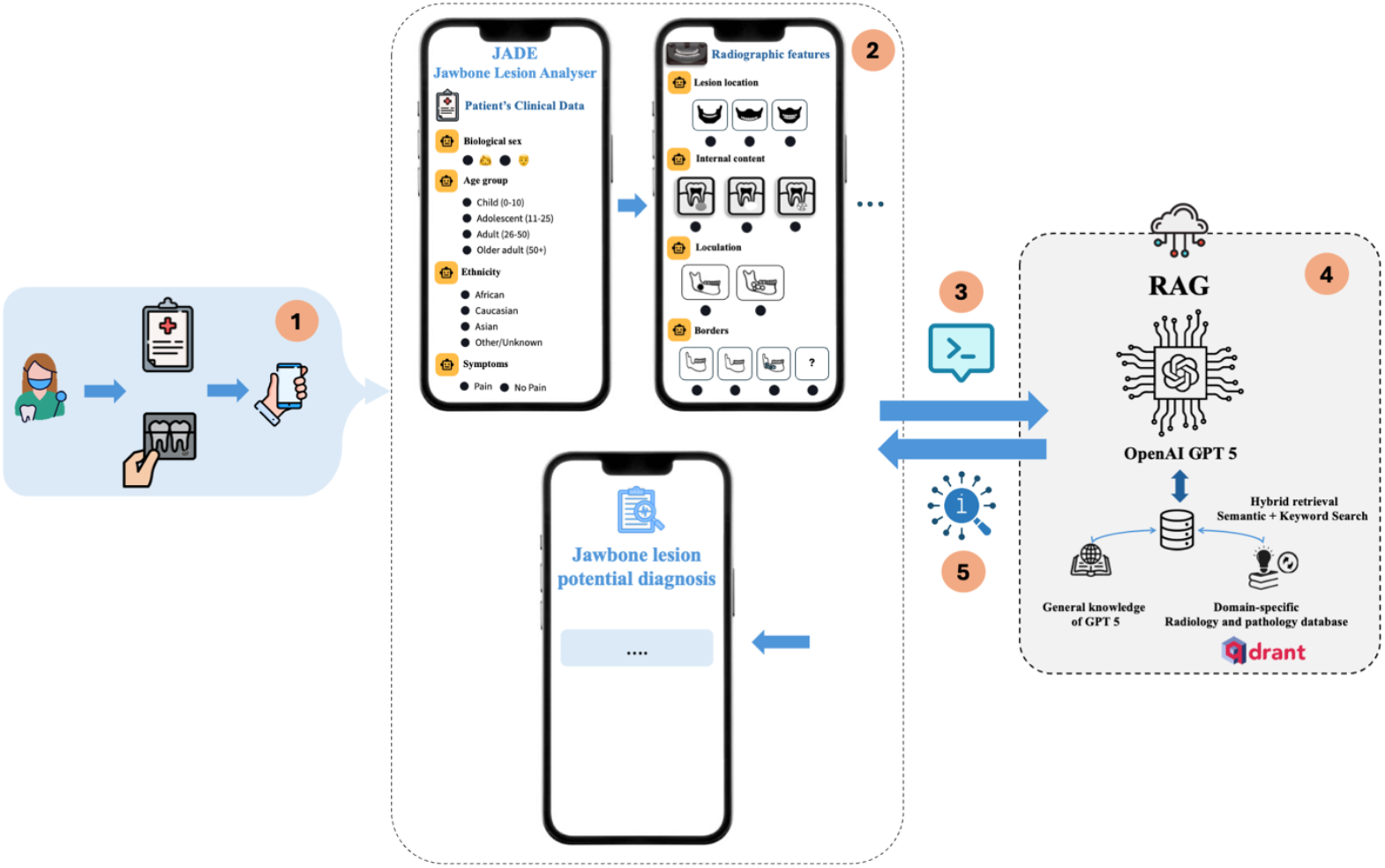
Overview of the JADE diagnostic workflow. (1) The clinician reviews the panoramic radiograph and the patient’s clinical data, then enters the radiographic characteristics of the jawbone lesion and relevant clinical information into the application. (2) The application interface organises these inputs into a structured and prioritised diagnostic query (questions). (3) A prompt is generated, including the reasoning format and instructions that define the diagnostic priority of each parameter. (4) The RAG module uses GPT-5 as the core LLM. The prompt is sent to the application engine, where GPT-5 retrieves information from the vector database using two sources: its internal general medical knowledge and the domain-specific radiology and pathology database hosted on Qdrant Cloud, accessed through a hybrid search. (5) The retrieved information is synthesised by GPT-5, converted into a human-readable diagnostic response, and sent back to the application interface for visualisation of the lesion diagnosis.

### Structured Query and Prompt

The diagnostic process starts with the clinician entering the answers to a set of ordered, predefined questions that record the clinical data of patients and the lesion’s radiographic characteristics through the application interface. These inputs are automatically arranged into a structured and prioritized query that forms the core of the model’s prompt (Table 1).

**Table 1.** Prioritized clinical and radiographic questions were used to structure the diagnostic prompt. The table outlines the set of standardized questions, their corresponding answer options, and their assigned priority levels, which collectively guide the system’s reasoning process in generating differential diagnoses.

| Question | Potential Answers | Priority |
| --- | --- | --- |
| Internal content | Radiolucent; Radiopaque; Mixed | High |
| Borders | Corticated; Well-defined (non-corticated); Diffuse | High |
| Loculation | Unilocular; Multilocular | High |
| Relation to surrounding teeth | At the apex of a vital tooth; At the apex of a non-vital tooth; In contact with / involving root; In contact with crown; Located in a missing-tooth area; Not related to any tooth | High |
| Jaw location | Mandible; Maxilla; Both | High |
| Cortical expansion | Yes; No | High |
| Root resorption | Yes; No | High |
| Number of lesions | 1; 2; $\geq 3$; Generalised | High |
| Gender | Male; Female | Medium |
| Age group | 0–10; 11–25; 26–50; >50 | Medium |
| Ethnicity | African; Caucasian; Asian; Other / Unknown | Medium |
| Regional location | Incisor region; Canine/Premolar region; Molar region; Ramus; TMJ; Maxillary sinus | Medium |
| Symptoms | Pain; No pain | Medium |
| Lesion size | <2 cm; 2–3 cm; >3 cm | Low |
| Lesion includes one or more teeth | Yes; No | Low |
| Tooth displacement or impaction | Yes; No | Low |

The reasoning step is guided using the following prompt format:

> *“You are an oral radiologist. Provide the differential diagnosis for a jawbone lesion based on the following clinical and radiographic characteristics*.*”*

The prompt also includes an instruction section that defines the diagnostic importance of each feature category. As indicated in Table 1:

- High-priority: Core radiograph characteristics that directly influence lesion classification.
- Medium-priority: Demographic and contextual factors that provide supportive diagnostic information.
- Low-priority modifiers: additional features that refine but do not determine the final diagnosis. This structured format ensures that the model considers the most clinically meaningful features first, as well as supporting information properly, and keeps a consistent diagnostic reasoning pathway across all cases.

### Core RAG Engine

The core engine of our proposed application is a RAG module built with GPT-5 as the language model. GPT-5 is an LLM pre-trained on a broad mixture of publicly available, licensed, and human-generated data, and further aligned using reinforcement learning from human feedback. All general medical knowledge used by the model is embedded within its parameters and does not rely on live web searches or external bases during inference.

The RAG module combines information retrieval with natural language generation using two complementary sources of information:

- General knowledge source: The reasoning capability of OpenAI’s GPT-5 (static knowledge cutoff: April 2025), which provides a broad contextual understanding and medical reasoning.
- Domain-specific knowledge source: A curated oral radiology and pathology database developed for this study by experienced oral and maxillofacial radiologists and pathologists. This database includes detailed descriptions of common jawbone lesions, their clinical manifestations, radiographic patterns, and differential diagnoses derived from authoritative reference works ^3,17–21^.

### Retrieval and Generation Workflow

To achieve efficient knowledge retrieval, the domain-specific dataset was embedded using text-embedding-3-large (chunk size = 1200, overlap = 200) and stored in a Qdrant Cloud vector database ^22^. JADE employs a hybrid retrieval approach that combines semantic (dense) and keyword-based (sparse) search to ensure both conceptual and keyword relevance.

Semantic (dense) retrieval uses embedding similarity to identify text chunks that are conceptually similar to the query, even when different terminology or phrasing is used. For example, a description of a “multilocular radiolucent mandibular lesion in a young adult” retrieves conceptually related cases such as ameloblastoma or odontogenic keratocyst.

Keyword (sparse) retrieval is based on BM25 ^23^, a classical information-retrieval algorithm that ranks documents according to how often a term appears and how informative or distinctive that term is within the dataset, while also accounting for document length. BM25 ensures that chunks containing explicitly mentioned terms (e.g., “mandible”, “multilocular”, “radiolucent”) are reliably captured.

Qdrant automatically merges dense and sparse results with its built-in hybrid score fusion, where both similarity signals are normalized and combined into a single relevance score. This default fusion logic ensures that neither semantic similarity nor exact keyword matching dominates, allowing conceptually aligned and lexically precise passages to be ranked appropriately. The top-ranked results are then provided to the RAG module, where GPT-5 integrates the retrieved evidence with its general reasoning capabilities to produce the most probable differential diagnosis. This workflow supports accurate, explainable, and reproducible diagnostic reasoning optimized for real-time clinical use.

### Hardware Setup

The application was developed in Python 3.12.10 and built using Streamlit ^24^, an open-source library for interactive web interfaces. To ensure reproducibility and environment portability, the application was containerized with Docker ^25^ and deployed via Google Cloud Run ^26^, allowing automatic scaling in response to usage demand.

For model development and testing, computations were performed locally on a workstation equipped with a 12th Gen Intel® Core™ i9-12900K processor operating at 3.20 GHz, 128 GB of RAM, and an NVIDIA RTX A5000 GPU with 24 GB of GDDR6 memory. The software environment ran on Microsoft Windows 11.

## Validation

### Data acquisition

The retrospective validation dataset consisted of 25 representative cases: radicular cyst (n = 3), fibrous dysplasia (n = 2), Stafne bone defect (n = 2), dentigerous cyst (n = 4), ameloblastoma (n = 2), cemento-osseous dysplasia (n = 3), central giant cell granuloma (n = 2), and simple bone cyst (n = 3). The cohort included 12 male and 13 female patients with a mean age of 39 years (range, 9–66 years). The majority of patients were Caucasian (n = 22), with one Asian patient and two Black patients.

Two oral radiologists and pathologists selected the cases based on their frequency and pathological relevance. Cases were selected if their panoramic radiographs presented at least one jawbone lesion without any artifacts or distortions affecting diagnostic evaluation.

All panoramic radiographs were obtained digitally using either the VistaPano S (Dürr Dental, Bietigheim-Hissingen, Germany; 2013–current; 73 kV, 12 mA, 7 s) or the Promax 2D (Planmeca, Helsinki, Finland; 2013–current; 68 kV, 10 mA, 16 s). All imaging was performed by licensed dental radiology technicians with at least three years of clinical experience.

All included cases were confirmed by either expert-based or histopathological diagnosis. In cases of disagreement, a mutual agreement was reached through discussion to ensure the accuracy and reliability of the dataset.

### Evaluation metrics

The diagnostic performance of the proposed system was evaluated and compared against several state-of-the-art LLMs, including GPT-5 standalone model, DeepSeek, Claude Sonnet 4, and Gemini 2.5 Flash. All models were evaluated based on their ability to generate a single most likely diagnosis, which was then compared with the reference diagnosis in the validation dataset.

#### Proportion of correct diagnosis

This metric represents the proportion of validation cases for which the model’s predicted diagnosis correctly matched the reference diagnosis.

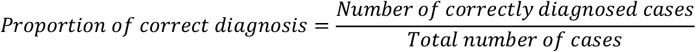

#### Majority agreement ratio (MAR)

MAR assesses intra-model stability by evaluating how consistent a model is in producing the same diagnosis across multiple independent runs for each case *i*.

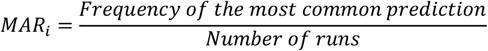

A higher MAR value means greater prediction consistency, regardless of whether the prediction is correct. Therefore, MAR reflects model stability, not diagnostic accuracy.

#### Mean stability

Overall stability of the model over the entire validation dataset.

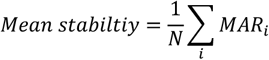

where *N*=25 validation cases.

### Statistical analysis

Diagnostic performance of the models was evaluated using Cochran’s Q test (Statsmodels, Python ^27^ ) to compare differences in binary outcomes (correct vs. incorrect diagnoses) across related cases. With five models included, the test was interpreted using the chi-square distribution with 4 degrees of freedom. If the test was significant, pairwise McNemar’s exact tests were calculated to identify specific differences between models. A Bonferroni correction was calculated to adjust for multiple comparisons, setting the family-wise significance level at *p* < 0.05 and the per-test threshold *p* < 0.0125 for the total of 4 pairwise comparisons. P-values falling between these two thresholds were considered statistically suggestive but not significant after correction. Each case was coded as 1 when the model’s prediction matched the ground truth diagnosis and 0 otherwise.

In addition, the mean ± standard deviation response time of the application was also calculated by recording the latency over 25 independent inference runs for each model.

## Results

The end-to-end diagnostic workflow of the JADE application is presented in Figure 2, showing how the system transforms structured patient data and lesion features into a final predicted diagnosis for a representative validation case.

**Figure 2.**
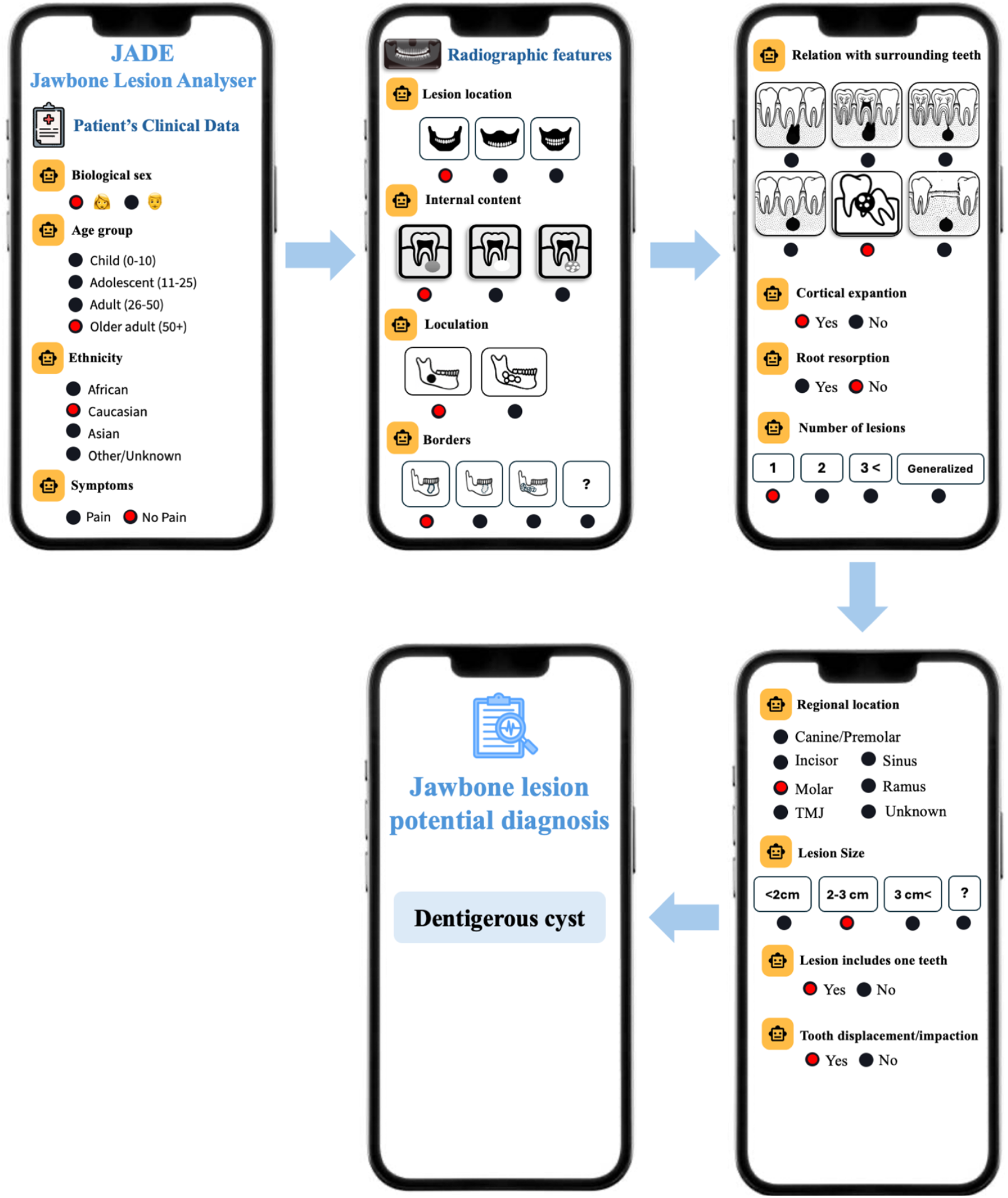
The figure presents how clinical and radiographic information is sequentially entered into JADE to generate a diagnosis for a sample case. In this example, the patient is an older adult (50 years +) female with no pain symptoms. The lesion is located in the mandibular molar region, linked to the crown, and presented as a single lesion. Radiographic features include a 2–3 cm size, central origin, and unilocular appearance, with corticated borders. The lesion involves multiple teeth, causes cortical expansion and tooth displacement, but no sign of root resorption. Based on these structured inputs, JADE correctly predicts a dentigerous cyst, matching the histopathological diagnosis.

Of all evaluated models, the proposed JADE system (RAG-based) showed the highest diagnostic accuracy, accurately identifying 20 out of 25 cases, outperforming all standalone LLMs (Figure 3). While GPT-5, which serves the core LLM of JADE without the RAG framework, could correctly identify only 10 cases, underscoring the added value of the RAG framework in contextual reasoning. Gemini 2.5 Flash showed the weakest performance, with only 9 correct diagnoses. On the other hand, Claude Sonnet 4.5 and DeepSeek-R1 showed a moderate performance with an almost 50/50 correct-to-missed ratio.

**Figure 3.**
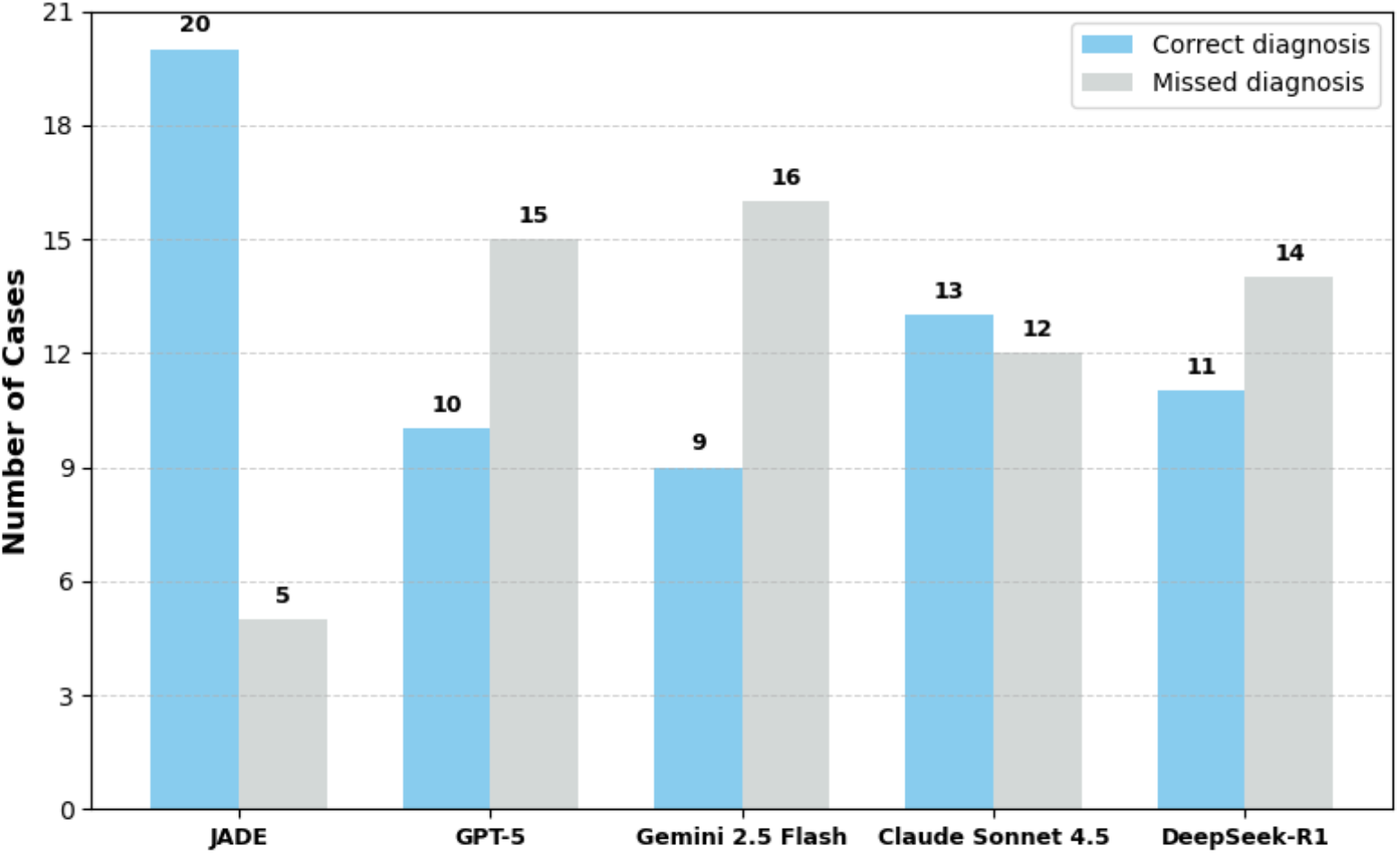
Comparative diagnostic performance of the proposed JADE system (RAG module) versus leading LLMs across 25 validation cases. JADE achieved the highest number of correct diagnoses, representing superior diagnostic accuracy, while the standalone GPT-5 model and other LLMs showed lower performance with more missed diagnoses.

The intra-model stability, measured by the majority agreement (mean ± SD), showed clear variability between the evaluated models (Figure 4). The proposed JADE system with RAG module demonstrated its efficiency in maintaining diagnostic stability of the system, achieving the highest mean value (0.90 ± 0.18) and generating uniform diagnostic predictions across repeated runs. Whereas standalone GPT-5 (0.64 ± 0.18) and Gemini 2.5 Flash (0.60 ± 0.18) presented the lowest stability with greater variability in diagnostic outputs. Claude Sonnet 4.5 and DeepSeek-R1 displayed moderate stability, positioned between the most and least stable models.

**Figure 4.**
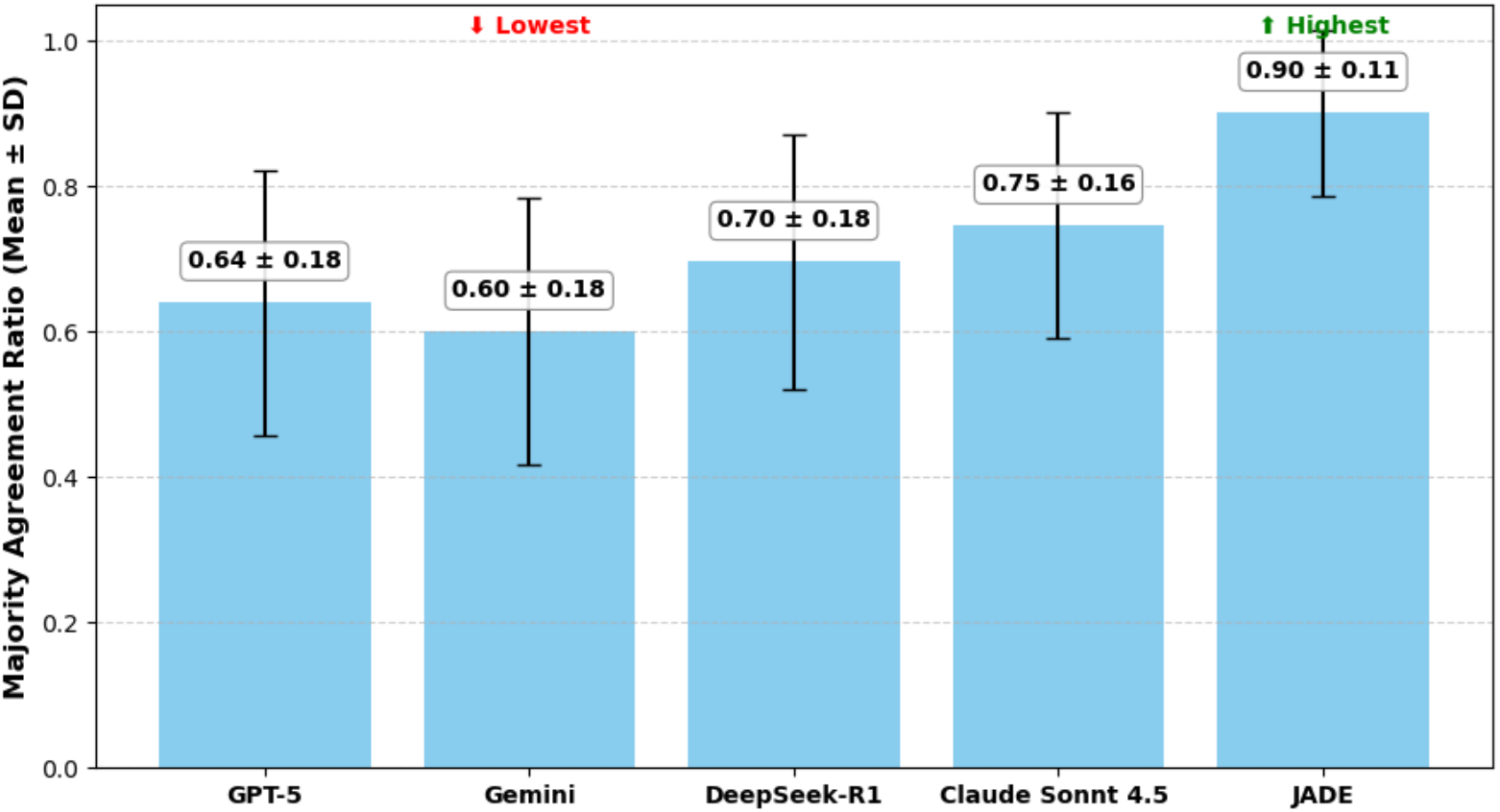
Comparison of intra-model stability of the proposed JADE system against the standalone LLMs, based on the MAR (mean ± SD). JADE achieved the highest stability, as indicated by the green highlights. In contrast, GPT-5 and Gemini 2.5 Flash, highlighted in red, showed the lowest stability among the evaluated models.

The statistical comparison of diagnostic performance showed significant differences across the evaluated models (Cochran’s Q = 33.2, *df* = 4, *p* < 0.001). Post-hoc McNemar’s exact tests with Bonferroni correction (family-wise α = 0.05; per-comparison α = 0.0125) showed that JADE significantly outperformed GPT-5, Gemini 2.5 Flash, and Claude Sonnet 4.5 with all corresponding p-values falling well below the adjusted significance threshold. In contrast, the comparison between JADE and DeepSeek-R1 did not reach statistical significance (*p* = 0.04) but remained statistically suggestive. This reflects the nature of McNemar’s test, which depends on discordant paired predictions rather than absolute accuracy gaps. JADE and Claude exhibited a more asymmetric pattern of disagreement (1 vs. 10), leading to a significant result, whereas the JADE–DeepSeek comparison showed fewer and less asymmetric discordant pairs (1 vs. 8), reducing statistical power and producing a non-significant, though suggestive, difference. Overall, these results indicate that the RAG-based system, JADE, generates substantially more accurate and consistent diagnostic decisions than the standalone LLMs evaluated.

Furthermore, the response time evaluation of the proposed JADE system showed its efficiency for real-time applications. The average response time of the JADE system, measured by recording the latency per case across ten independent runs, ranged from 5 ± 0.4 seconds and 8 ± 0.6 seconds, with a mean latency of 6 ± 0.5 seconds.

Overall, JADE outperformed standalone LLMs in diagnostic accuracy and consistency, achieving superior diagnostic accuracy and near real-time performance. These findings support the feasibility and clinical potential of RAG frameworks for diagnostic decision support in oral radiology.

## Discussion

This study introduces JADE, a novel RAG diagnostic framework designed to support and enhance diagnostic reasoning for jawbone lesions. Instead of relying only on the general knowledge embedded within GTP-5 or exclusively on the domain-specific references, JADE includes both sources through a RAG pipeline. Using a hybrid search strategy (keyword-based and semantic retrieval), the system retrieves validated radiologic and pathologic information from an expert-curated jawbone lesion dataset and then synthesizes this information with internal medical reasoning of GPT-5 to produce clinically coherent, context-appropriate differential diagnoses. This integration of general and expert-validated knowledge allows JADE to generate diagnostic predictions that are both medically related and case-specific, overcoming the limitations of standalone LLMs that rely on patterns learned during training. Moreover, JADE was developed as a cloud-based, mobile-adapted application that makes it accessible and user-friendly for general dentists, oral radiologists, and students in dental, radiological, and educational settings.

The RAG pipeline with a hybrid retrieval strategy allows JADE to more reliably identify lesion-specific radiographic and clinical patterns that standalone LLMs often overlook. Its high intra-model stability also shows that the predictions of the proposed system are less influenced by stochastic variation over repeated prediction turns, a challenge that LLMs have due to relying only on internal training representations. Such stability makes the proposed system clinically relevant since reproducible outputs limit ambiguity and strengthen confidence in the AI-assisted decision-making process.

The statistical analyses further support the reliability of JADE’ diagnostic power by showing that its improvements were not because of random variations but due to systemic differences in performance. The overall significance of the evaluated models shows that retrieval augmentation considerably enhances diagnostic accuracy. Post-hoc comparisons further highlighted that JADE outperformed commonly used standalone LLMs such as GPT-5, Gemini 2.5 Flash, and Claude Sonnet 4.5, showing the added value of integrating domain-specific knowledge and a hybrid retrieval approach. Although JADE achieved higher accuracy than both Claude and DeepSeek, only the comparison with Claude remained statistically significant after correction. It is also worth noting that the low consistency of the standalone LLM highlights the limitations of prompt-only reasoning in the specialized diagnostic domain.

In addition, JADE achieved a mean end-to-end response time of 6 seconds. This includes both the hybrid retrieval step (which adds only a minor overhead of <0.5 seconds) and GPT-5 reasoning. The overall latency is comparable to, and in some cases faster than, standalone LLMs’ responses (typically 4–6 seconds), indicating that the addition of the RAG component does not substantially increase response time. As a mobile-accessible cloud platform, JADE therefore operates within the range required for real-time clinical decision support and educational use.

These findings are aligned with recent studies evaluating LLM models in oral diagnosis. Kaygisiz et al. ^8^ reported that DeepSeek-v3 showed moderate diagnostic accuracy and performed better than GPT-4o, with scores of 4.02 ± 0.36 versus 3.15 ± 0.4, and outperformed GPT-4o in 9 of 16 simulated cases However, both models showed limited real-world applicability, as in this study they failed to correctly diagnose almost half of the representative cases, highlighting the limitations faced by standalone LLMs in complex diagnostic scenarios. Hassanein et al.^28^ found Top-1 accuracy on 80 clinical cases, with DeepSeek-V3 achieving 45% and GPT-4o 40% and only reaching 70% accuracy at the Top-3. Zhuang et al. ^29^ further reported diagnostic accuracy of 71% for Claude sonnet 3.5 and 57% for GPT-4o, consistent with our finding in which Claud sonnet outperformed GPT-based models. In another study, Pradhan et al. ^30^ found that GPT-4o correctly diagnosed 28 out of 42 potentially malignant lesions. Whereas Gemini demonstrated substantially weaker performance with only 15 correct diagnoses. Similarly, Kim et al.^7^ showed that ChatGPT-4 achieved a concordance rate of 41.4%, comparable to clinicians (43.2%) and slightly below the ORAD system (45.6%). Overall, these results confirmed the inconsistent and often limited diagnostic reliability of standalone LLMs, a pattern also reflected in our findings and underscoring the added value of retrieval-augmented systems such as JADE.

Despite the improvements achieved in the diagnostic accuracy of the proposed system, several cases were still misdiagnosed. This indicates that the RAG mechanism and hybrid retrieval strategy alone cannot fully eliminate diagnostic errors, particularly when the information provided in the structured questionnaire is incomplete or lacks key discriminative clinical or radiographic features. This is particularly important when the possible diagnoses are very similar, where targeted and lesion-specific questions can help guide the right retrieval and support more accurate differentiation. Future improvements for the proposed system can focus on an agentic self-evaluating mechanism able to assess the predicted outcome against the key clinical and radiographic parameters provided in the query. This pipeline could automatically perform re-evaluation or refinement of the prompt when the predicted diagnosis is consistent with the described lesion characteristics. In addition, the domain-specific database could also be optimised by reconstructing it into shorter, parameter-specific documents to allow more precise retrieval that can be dynamically controlled by an agentic mechanism. Another limitation of this study is the relatively small validation dataset that limits the generalisability of the system. Future studies should overcome these challenges and prioritize expanding the dataset to include a wider range of jawbone lesions.

## Conclusion

This study introduces JADE, a retrieval-augmented AI-assisted diagnostic system, as a proof-of-concept for the diagnosis of jawbone lesions. Through integrating GPT-5 general reasoning power with structured domain-specific hybrid retrieval, the proposed system shortened the gap between LLMs and clinically based diagnostic predictions. Developed as a cloud-based, mobile-adapted tool, it provides real-time interaction and accessibility for both clinical and educational use. Although further improvements are required to improve the generalisability, the postposed system demonstrated strong potential to turn into a practical diagnostic support tool for jawbone lesions. With continued development and wider validation, this pipeline may contribute to more accurate, consistent, and accessible diagnostic assistance within oral and maxillofacial radiology.

## Data Availability

The data supporting the findings of this study are not publicly available due to ethical and privacy restrictions but are available from the corresponding author upon reasonable request.

